# Intestinal mucosal immune responses to novel oral poliovirus vaccine type 2 in newborns

**DOI:** 10.1101/2025.03.05.25323405

**Authors:** Audrey Godin, Elizabeth B. Brickley, Ruth I Connor, Wendy F. Wieland-Alter, Joshua A. Weiner, Margaret E. Ackerman, John F. Modlin, Omar M. Sajjad, Minetaro Arita, Chris Gast, Bernardo A. Mainou, Khalequ Zaman, Masuma Hoque, Sohel Rana, Ananda S. Bandyopadhyay, Peter F. Wright

**Affiliations:** Health Equity Action Lab, Department of Infectious Disease Epidemiology and International Health, London School of Hygiene & Tropical Medicine, London, United Kingdom; Department of Pediatrics, Geisel School of Medicine at Dartmouth, Dartmouth Health, Lebanon, New Hampshire, USA; Department of Microbiology and Immunology, Geisel School of Medicine at Dartmouth, Hanover, New Hampshire, USA; Thayer School of Engineering, Dartmouth College, Hanover, New Hampshire, USA; Department of Virology II, National Institute of Infectious Diseases, Tokyo, Japan; PATH Center for Vaccine Innovation and Access, Seattle, Washington, USA; Division of Viral Diseases, US Centers for Disease Control and Prevention, Atlanta, Georgia, USA; International Centre for Diarrhoeal Disease Research, Bangladesh; Bill & Melinda Gates Foundation, Seattle, Washington, USA

## Abstract

**Background:** Approximately 1.2 billion doses of novel type 2 oral polio vaccine (nOPV2) have been administered in response to circulating vaccine-derived poliovirus type 2 (cVDPV2) outbreaks since 2021. Although infants are eligible to receive the vaccine from birth, the induction of intestinal mucosal immunity by nOPV2 in newborns has not been directly evaluated.

**Methods:** In a randomized, placebo-controlled, phase 2 clinical trial in Bangladesh (2020–2021), 215 healthy newborns received two doses of either nOPV2 (n=110) or placebo (sucrose; n=105), at birth (0-3 days) and 4 weeks later. Intestinal mucosal immunity was assessed by measuring poliovirus type 2 (PV2)-specific neutralizing activity and immunoglobulin (Ig)A levels in stool collected biweekly from birth to 8-weeks.

**Results:** Newborns vaccinated with two doses of nOPV2 had strong intestinal mucosal immune responses that differed significantly from the placebo group (p<0.0001 for PV2-specific neutralization from 2 weeks onward and p≤0.007 for PV2-specific IgA from 4 weeks onwards). Positive PV2-specific neutralization in stool (i.e., titers ≥16) was detected in 51.8% (57/110) of nOPV2-vaccinated newborns at 4 weeks and 90.0% (99/110) at 8 weeks (4 weeks after the second dose). Notably, PV2-specific antibody titers following the second dose were very similar for newborns who did and did not have first dose responses (p=0.67 for neutralization and p=0.38 for IgA at 8 weeks).

**Conclusions:** Vaccination with two doses of nOPV2 in neonates induced high intestinal mucosal immune responses. In cVDPV2 outbreak settings, neonatal administration of nOPV2 may be a strategy to enhance population-level intestinal mucosal immunity.

**Trial registration number** NCT04693286

## Introduction

Circulating vaccine-derived type 2 polioviruses (cVDPV2s) are currently the leading cause of poliomyelitis and threaten global poliovirus (PV) eradication efforts (1). In response to cVDPV2 outbreaks, approximately 1.2 billion doses of novel type 2 oral poliovirus vaccine (nOPV2), which is less likely to acquire genetic mutations linked to neurovirulence than the historically used Sabin oral poliovirus vaccine type 2, have been administered since 2021 (2–4). In infants (18-22 weeks), children (1-5 years), and adults (18-50 years), nOPV2 has been shown to be safe, well-tolerated, and capable of inducing both serum immunity, which confers individual protection against paralytic poliomyelitis, and intestinal mucosal immunity, which reduces poliovirus replication and onward fecal-oral transmission (5–13).

Although infants are eligible to receive nOPV2 from birth, the induction of intestinal mucosal immunity by nOPV2 in newborns has not been directly evaluated. Using paired stool and serum samples from 215 healthy newborns participating in a phase 2 clinical trial of nOPV2 in Bangladesh (9), we evaluated the induction of intestinal mucosal immunity (i.e., PV type 2 (PV2)-specific neutralizing activity and immunoglobulin [Ig] A levels in stool) in newborns vaccinated with two doses of nOPV2 (at birth and 4 weeks of age) compared to a placebo arm and investigated the influence of maternally-derived antibodies in the detection of intestinal mucosal responses during the neonatal period.

## Methods

### Study design and participants

Intestinal mucosal immunity was measured after one and two doses of nOPV2 in newborns included in a placebo-controlled, randomized, double-blinded phase 2 clinical trial (NCT04693286; N=330) conducted at the Matlab Health Research Center, Chandpur, Bangladesh, between September 2020 and August 2021. The study design, eligibility criteria, interventions, primary safety, and serum immunogenicity outcomes have been reported previously (9). Eligible participants included healthy, singleton newborns born at ≥ 37 weeks gestation. The main exclusion criteria were: (i) diagnosis or suspicion of immunodeficiency disorder in the newborn or immediate family member, (ii) contraindication for venipuncture, (iii) infection or illness requiring hospitalization, (iv) vomiting or intolerance to liquids <24 hours before enrollment, and/or (v) prior receipt of any poliovirus and/or rotavirus vaccine.

Participants received two doses of nOPV2 (10^5.0+/-0.5^ 50% cell culture infectious dose (CCID_50_), PT Bio Farma, Bandung, Indonesia) or placebo (sucrose in Basal Medium Eagle and buffer, PT Bio Farma) as two oral drops (0.1mL) at birth (0-3 days) and 4 weeks later. Serum was collected at birth, weeks 4 and 8; stool was collected every two weeks from birth. Samples were shipped frozen to the Polio and Picornavirus Branch of the Division of Viral Diseases, Centers for Disease Control and Prevention (CDC, Atlanta, GA, USA), where serum samples were analyzed using a PV serotype-specific microneutralization assay (PMID 26983734) and stool samples were qualitatively evaluated for the presence of PV serotype-specific shedding using real-time reverse transcription polymerase chain reaction (RT-PCR) (9).

### Laboratory Procedures

Intestinal mucosal immunity was evaluated in stool samples from a randomly selected subset of participants (110 from the vaccine arm and 105 from the placebo arm). Samples were aliquoted and sent frozen to the Geisel School of Medicine at Dartmouth College (Hanover, NH, USA). PV1-, PV2-, and PV3-specific neutralization titers in stool were reported as the reciprocal of the limiting dilution needed to achieve 60% neutralization of luciferase-expressing wild-type-derived polio pseudo-viruses (14). Neutralization titers <4 (i.e., lower limit of detection) were recorded as 2. In the nOPV2 group, samples with PV2-specific titers >512 were serially diluted further, up to 16,384, to capture the highest end-point titers. Total IgA concentrations (µg/ml) and PV1-, PV2-, PV3-specific IgA and IgG levels (expressed as median fluorescence intensity [MFI] and, if negative after background signal subtraction, recorded as half of the lowest observed homotypic MFI) in stool were quantified using a multiplex assay developed by coupling monovalent inactivated poliovirus vaccines to fluorescently coded magnetic microspheres, as described previously (15). Lactoferrin (ng/ml) in stool was measured using a commercially available enzyme-linked immunosorbent assay (ELISA) per manufacturer’s instructions (Lactoferrin Scan^®^ TechLab, Inc., Blacksburg, VA, USA).

### Statistical analyses

Differences in the distribution of PV serotype-specific neutralization, IgA, and IgG levels were assessed using Mann-Whitney U tests. Differences in the proportion of positive neutralizing activity (defined as neutralization titers ≥16) and detectable vaccine viral shedding (PV2-positive RT-PCR) in stool at each visit and/or ever (i.e., in at least one visit) were assessed using Pearson’s chi-squared tests. To evaluate the effect of the second dose of nOPV2 on intestinal mucosal immunity, we categorized participants as birth dose responders (neutralization titers in stool ≥16 at 2 and/or 4 weeks [before the second dose]) or non-responders (titers <16 at 2 and 4 weeks). To investigate if placentally-transferred maternal antibodies interfered with the detection of intestinal mucosal responses after the birth dose, we calculated the odds ratios for the association of (i) positive neutralization and (ii) PV2-positive RT-PCR in stool at 2 and/or 4 weeks with the magnitude of serum neutralization at birth in tertiles. As a sensitivity analysis for the threshold of positive neutralization, we repeated the analyses using titers ≥4 (i.e., detectable). As a posthoc analysis to determine whether the IgA levels observed in stool at baseline (0-3 days after birth) might be explained by breastfeeding, we investigated the relationship between lactoferrin, total IgA, and PV serotype-specific IgA levels in stool at baseline using Spearman’s rank correlations.

All *P* values are from 2-sided tests. All analyses were performed using Stata, v17.0, and R, v4.2.0.

## Ethics

Research and ethical review committees at the International Centre for Diarrhoeal Disease Research, Bangladesh provided ethical approval. Written informed consent was provided by all newborns’ parents/guardians before enrollment, including use of samples for further polio-related studies. Dartmouth-Hitchcock Institutional Review Board approved the de-identified samples analysis (#02002410).

## Results

We evaluated PV1, PV2, and PV3-specific intestinal mucosal immunity in 215 newborns (50.2% female), representing 65% of the participants in the parent trial. At baseline, infants in the nOPV2 and placebo groups were similar in terms of sex, age, birth weight, breastfeeding status, and Bacillus Calmette-Guérin (BCG) vaccination (Table S1). Most newborns were reported to be exclusively breastfed at birth (99.5% [214/215]) and at least partially breastfed at 4 weeks (99.1% [213/215]). The median (interquartile range, IQR) total IgA measured in stools was 0.5 (0.2-35.1) µg/ml at baseline and ranged from 46.0 (27.8-75.0) to 61.3 (39.8-92.8) µg/ml during subsequent visits. At baseline, positive neutralizing activity (i.e., titers ≥16) in stool was detected in 1.8% (2/110) participants in the nOPV2 group and 1.9% (2/105) in the placebo group (median [IQR] titers: 2 [2-2]).

The nOPV2 birth dose induced positive neutralizing activity in stool in 28.2% (31/110) participants by 2 weeks (median [IQR] titers: 2 [2-23.4]) and 51.8% (57/110) at 4 weeks (median [IQR] titers: 22.8 [2- 284.3]); compared with 4.8% (5/105) and 1.9% (2/105) at weeks 2 and 4 respectively (median [IQR] titers: 2 [2-2]) in the placebo group (Table 1, Figure 1). PV2-specific IgA levels in stool rose progressively after the nOPV2 birth dose to a median (IQR) MFI of 711.6 (228.6-1599.6) at 4 weeks (Table 1, Figure 2). In contrast, PV2-specific IgA levels in the placebo group and PV1- and PV3-specific IgA levels in all participants remained low (Figure 2). Stool PV2-specific IgG levels were largely undetectable in follow-up (Table 1).

**Figure 1.**
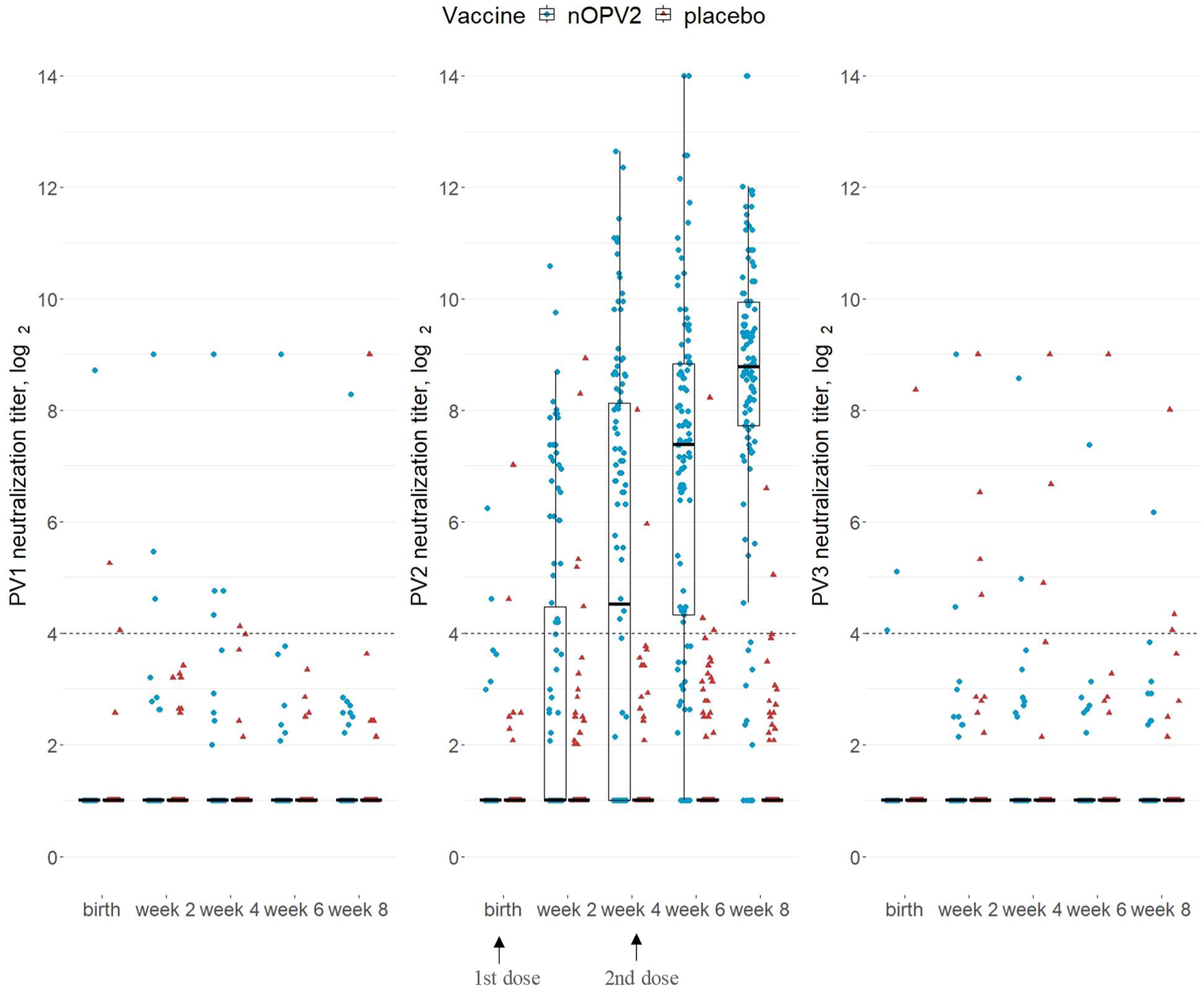
Distribution of poliovirus serotype-specific (PV1, PV2, PV3) log_2_ neutralization titers in stool after vaccination with the novel oral polio vaccine type 2 (nOPV2) or placebo. Colors and shapes indicate the vaccine received. Horizontal lines from the boxplots indicate the median; box, and whisker indicate the interquartile range (IQR) and 1.5*IQR. The dashed lines indicate the limit of positivity (i.e., titers ≥16). Neutralization titers <4 (i.e., lower limit of detection) were recorded as 2 (i.e., log_2_ titers=1). The upper limit of detection of neutralization titers was set at 512 (i.e., log_2_ titers=9) for PV1 and PV3 and 16384 (i.e., log_2_ titers=14) for PV2.

**Figure 2.**
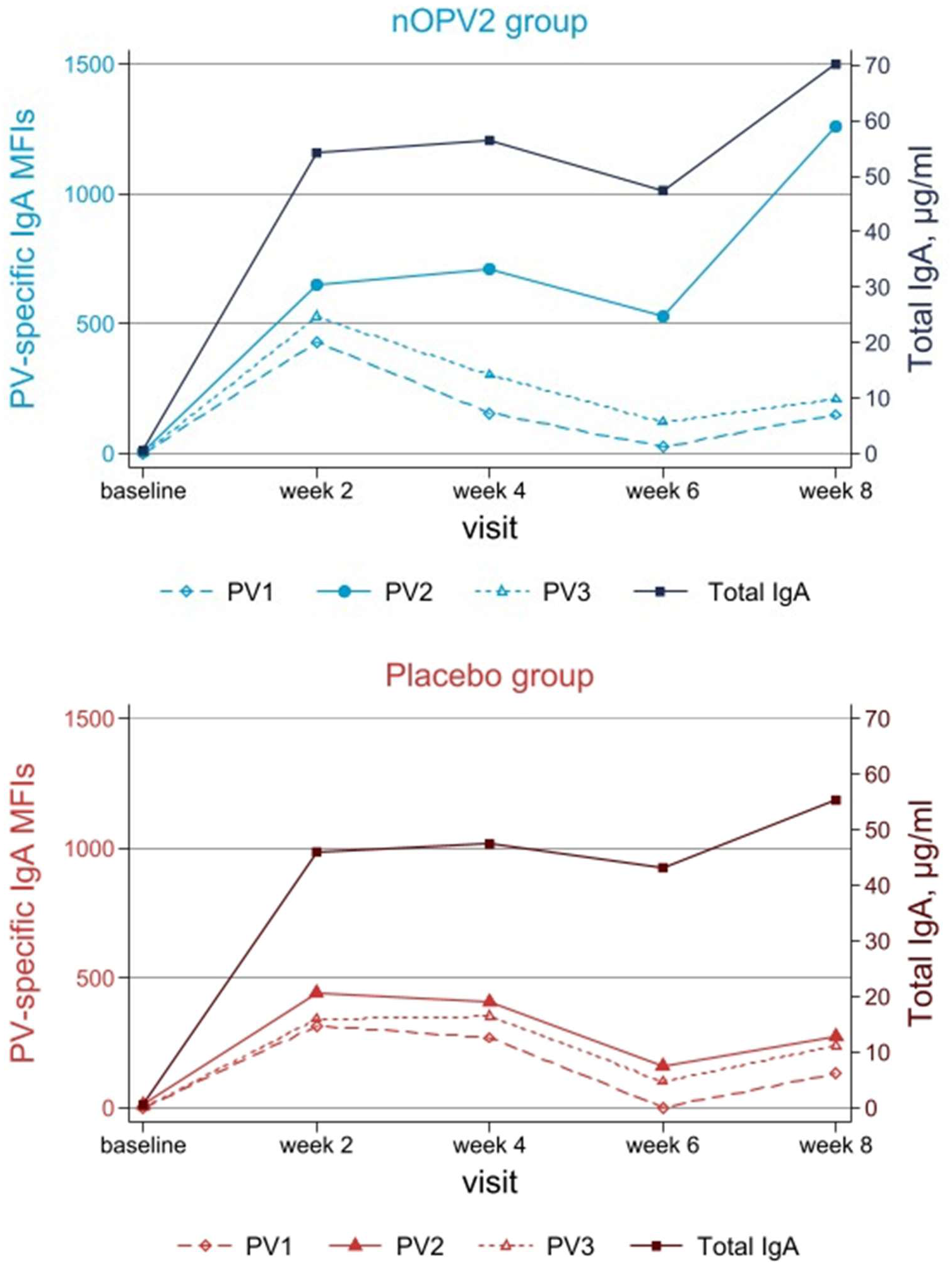
Median total immunoglobulin A (IgA, µg/ml) and poliovirus serotype-specific (PV1, PV2, PV3) median fluorescence intensities (MFIs) IgA in stool after vaccination with the novel oral polio vaccine type 2 (nOPV2) or placebo.

**Table 1.**
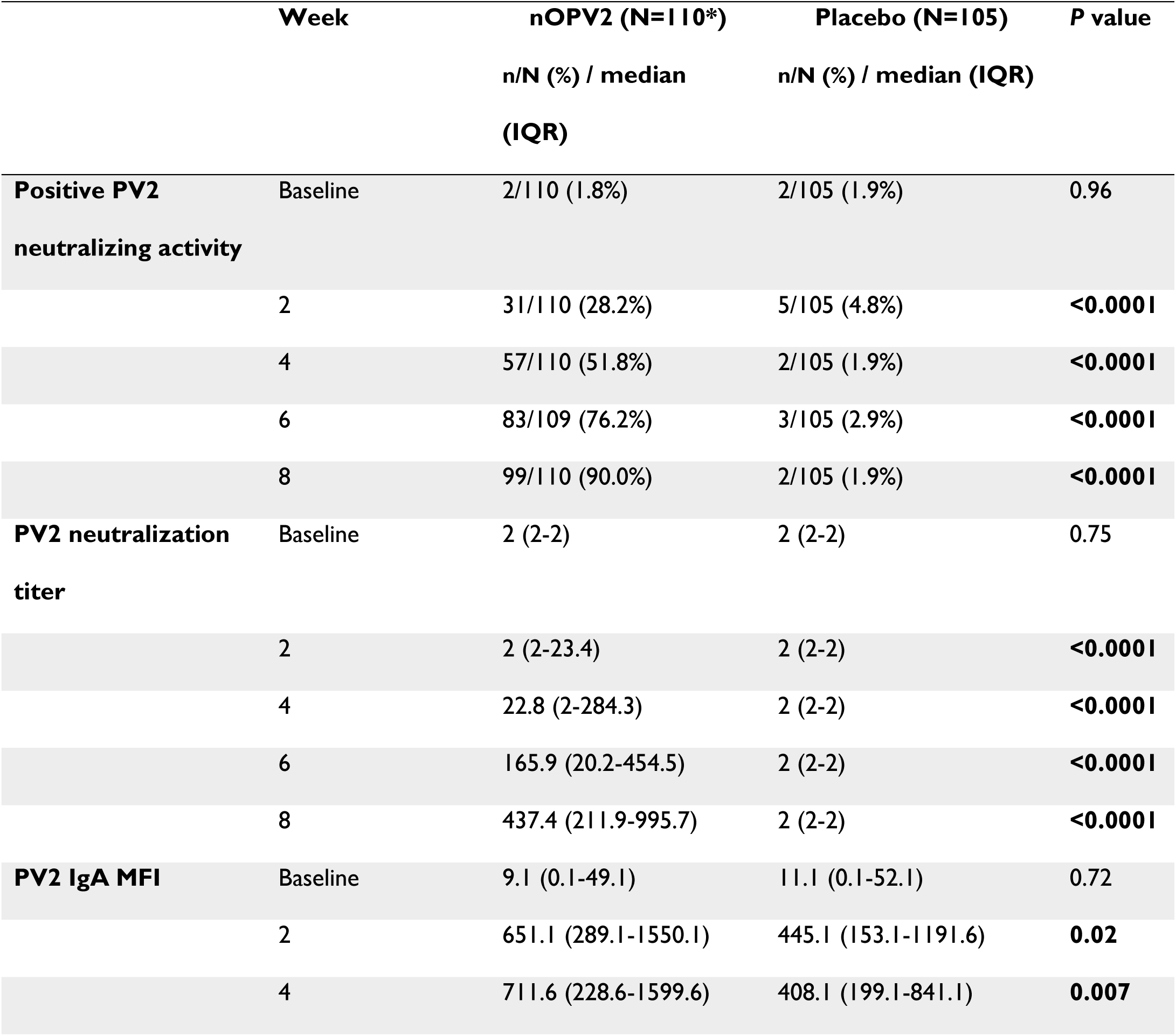
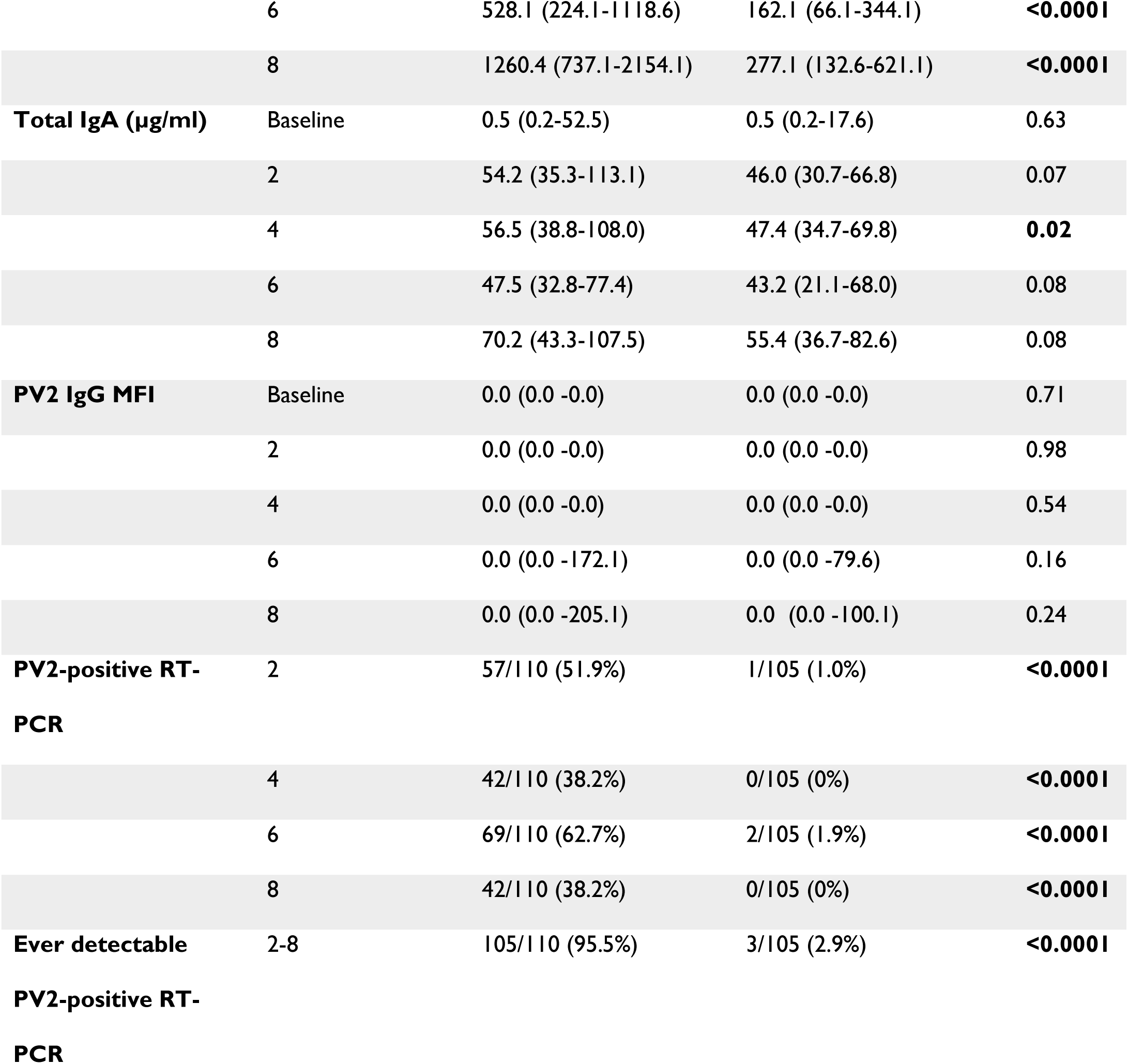
Poliovirus type 2 (PV2)–specific intestinal mucosal responses following administration of novel type 2 oral poliovirus vaccine (nOPV2) or placebo. *P* values are from Pearson’s chi-squared or Mann-Whitney U tests. Neutralization titers ≥16 were considered a positive neutralizing activity. Abbreviations: IgA, Immunoglobulin A; MFI, median fluorescence intensity; IQR, interquartile range; RT-PCR, reverse transcription polymerase chain reaction. *n=109 at baseline for total IgA, in week 4 for the PV2 IgA, and in week 6 for all measurements in the nOPV2 group.

We compared the 59 (53.6%) nOPV2 birth dose responders (i.e., with PV2-specific neutralization titers ≥16 at 2 and/or 4 weeks) to the 51 (46.4%) birth dose non-responders. At 2 weeks, 93.2% (55/59) of responders had PCR-detectable PV2 shedding in stool compared with 3.9% (2/51) of non-responders (Table 2). In contrast, 92.2% (47/51) of the non-responders had detectable PV2 shedding in stool 2 weeks after the second dose. Among birth dose responders, 37.3% remained RT-PCR PV2-positive at 6 weeks reflecting either virus shedding from the second dose or residual shedding from their birth dose. Most birth dose non-responders (90.2% [46/51]) had positive PV2-specific neutralization in stool at 8 weeks (i.e., 4 weeks after the second dose). Notably, at 8 weeks PV2-specific neutralization titers and IgA levels in stool were comparable between the birth dose responders and non-responders (p≥0.38, Table 2, Figure S1).

**Table 2.**
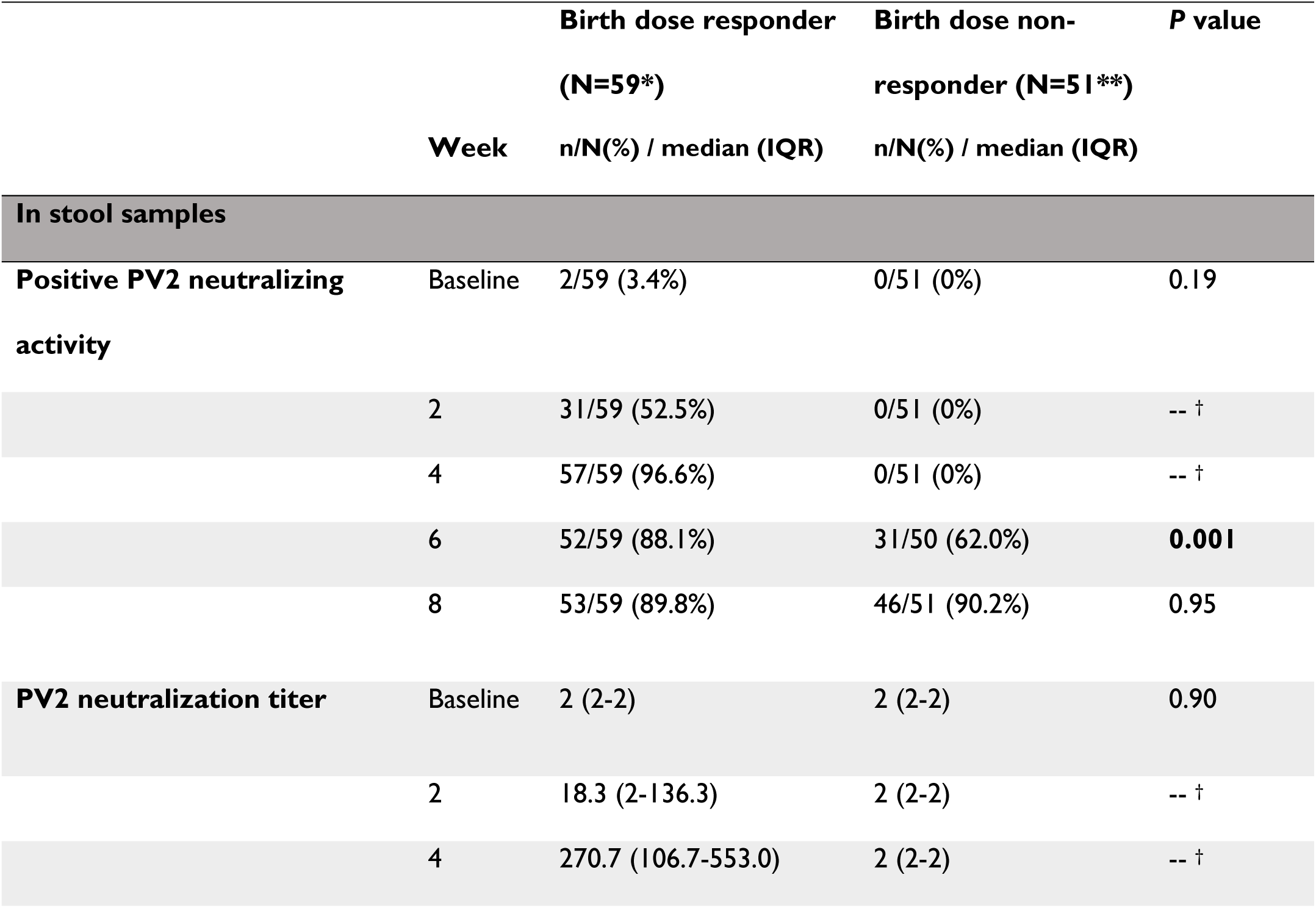
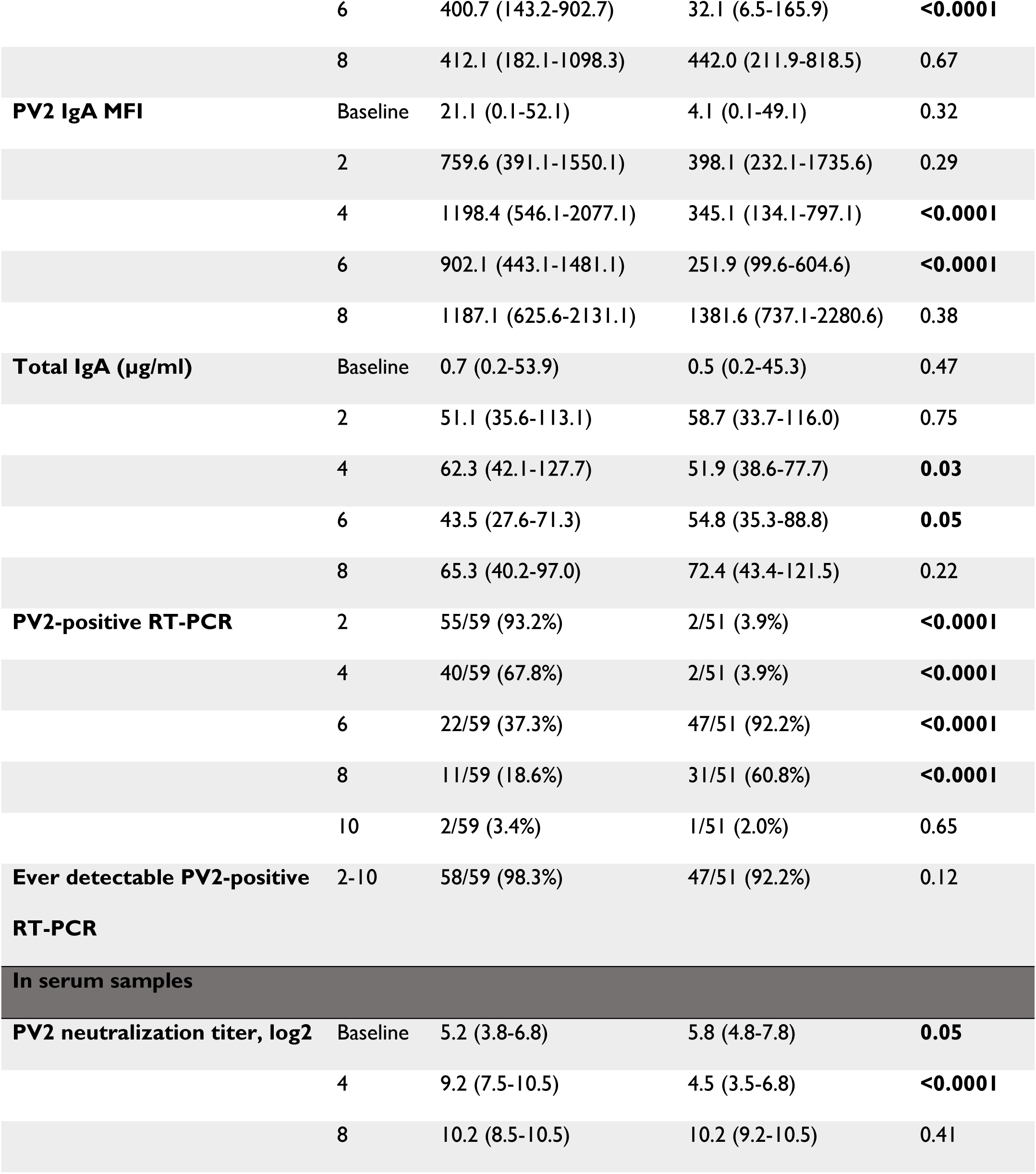
Stool and serum poliovirus type 2 (PV2)–specific responses following administration of the novel type 2 oral poliovirus vaccine (nOPV2) second dose in responders or non-responders to the first dose. *P* values are from Pearson’s chi-squared or Mann-Whitney U tests. Responders are participants with stool neutralization titers ≥16 (i.e., positive neutralizing activity) at week 2 and/or week 4. Abbreviations: IgA, Immunoglobulin A; MFI, median fluorescence intensity; IQR, interquartile range; RT-PCR, reverse transcription polymerase chain reaction. *n=58 at week 4 for IgA and at baseline for total IgA in stool; at week 8 for neutralization in serum **n=50 at week 6 for all measurements in stool; at week 8 for neutralization in serum ^†^ No p-values were calculated as the groups were defined by the absence of neutralizing activity at weeks 2 and 4.

Birth dose responders and non-responders did not differ in terms of baseline characteristics of sex, weight, breastfeeding, and BCG receipt (Table S2). However, 41.5% (17/41) of the newborns who had low serum neutralization titers at baseline (i.e., lowest tertile) had positive neutralizing activity in stool 2 weeks after receiving nOPV2 as compared to 20.0% (7/35) of those with high baseline serum neutralization (i.e., highest tertile, Table 3). Relative to newborns with high serum neutralization titers at baseline, those with low serum neutralization titers had 2.8 (95% CI 1.0-8.0, p=0.05) times the odds of positive neutralizing activity in stool and 2.6 (95% CI 1.0-6.5, p=0.05) times the odds of being RT-PCR PV2-positive at 2 weeks (Table 3). There was no association between baseline serum neutralization titers and positive neutralizing activity in stool at 4 weeks. In sensitivity analysis using the limit of detection of stool neutralization (titers ≥4), the odds ratio of the association of detectable stool neutralization at 2 weeks and low baseline serum neutralization was 3.9 (95%CI 1.4-10.6, p=0.008; Table S3).

**Table 3.**
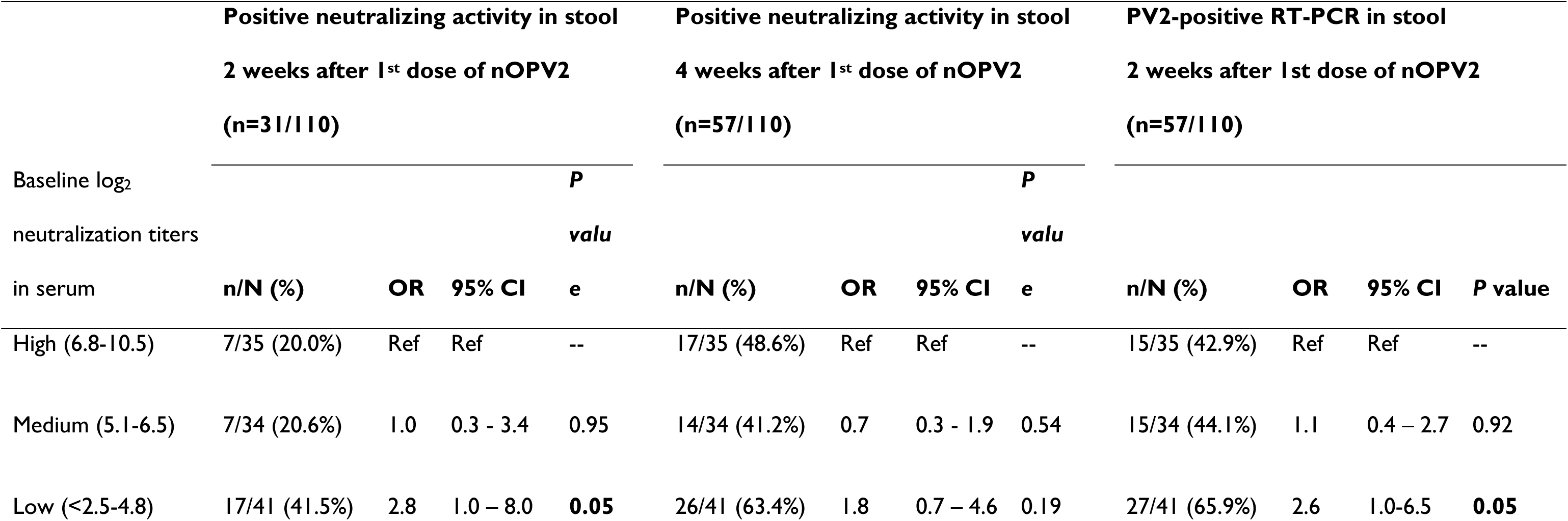
Association of poliovirus type 2 (PV2)-specific positive neutralizing activity and PV2-positive RT-PCR in stool two and four weeks after receiving the first dose of novel type 2 oral poliovirus vaccine (nOPV2) with the magnitude of PV2-specific serum neutralization titers at baseline (in tertiles). *P* values are from Pearson’s chi-squared tests. Neutralization titers ≥16 were considered a positive neutralizing activity. Abbreviation: RT-PCR, reverse transcription polymerase chain reaction; OR, odds ratio; 95% CI, 95% confidence interval.

By 8 weeks (i.e., 4 weeks after the second dose), 90.0% (99/110) of all nOPV2 recipients had positive PV2- specific neutralizing activity in stool (median [IQR] titers: 437.4 [211.9-995.7], Table 1). In contrast, few newborns had positive PV1- or PV3-specific neutralizing activity in stool during follow-up, indicating little circulation of the vaccine virus or strain cross-reactivity in the assay (Figure 1, Table S4). In sensitivity analysis using the limit of detection of neutralization (titers ≥4), 106/110 (96.4%) participants in the nOPV2 group and 17/105 (16.2%) in the placebo group had detectable neutralizing activity by 8 weeks (Table S5). Overall, the PV2-specific neutralization titers and IgA MFIs in stool in the nOPV2 group were similar between males and females and between the participants who did and did not receive a BCG vaccine at birth (Table S6).

As a posthoc analysis, we investigated whether the low levels of IgA in stool observed in both the vaccine and placebo arms at baseline (Table 1) were associated with lactoferrin (as a biomarker of breastmilk consumption). We observed strong positive correlations between total IgA, PV serotype-specific IgA MFIs, and lactoferrin concentrations in the newborns’ baseline stool samples (Spearman’s rho ≥0.41, p<0.0001 for all, Figure S2).

## Discussion

Our findings provide evidence that vaccination with two doses of nOPV2, administered at birth and 4 weeks, induces strong PV2-specific intestinal mucosal immunity as measured by functional neutralization and binding antibodies in stool. Although only half of the vaccine recipients had functional neutralizing activity after the birth dose, 90% of all nOPV2 recipients had positive PV2-specific neutralizing activity in stool following the second dose. Overall, our findings support the WHO recommendation that nOPV2 may be used in cVDPV2 outbreak responses from birth (16); however, a second dose is necessary to effectively elicit intestinal mucosal immunity in most newborns.

Although it has been reported that neonatal immunization with some vaccines induces weaker and shorter serum immune responses (17,18), the median PV2-specific neutralization titers in stool observed after two doses of nOPV2 (median of 437 at 8 weeks) were similar to the peak neutralization titers reported in OPV2-naïve infants (28 weeks of age) in Chile after one dose of mOPV2 (median ranging from 362 to 388 across the trial’s randomization arms) (19) and higher than the peak neutralization titers reported in OPV2-naïve toddlers (18 months old) in Panama after one dose of nOPV2 (median of 84) and OPV2-naïve adults in Belgium after one dose of nOPV2 (median of 2) (8,20). Large variability in the serum immunogenicity after an OPV birth dose has been observed (reviewed in 16), and further research is needed to determine whether the difference in the magnitude of neutralization responses in stool is due to maternal immunity, the age of vaccine recipients, the number of doses administered, or the vaccine setting.

The PV2-specific neutralization in stools was also slower to peak after receipt of nOPV2 at both birth and 4 weeks than previously observed in older individuals. We have previously reported that PV2-specific neutralization and IgA MFIs in stool peaked by 2 weeks after mOPV2 vaccination in OPV2-naïve infants (aged 18-32 weeks) in Latin America (19,22). Only 38% of nOPV2 recipients in our study had detectable neutralization (titers>4) 2 weeks after the birth dose; in contrast, we had previously observed detectable PV2-specific neutralization in stool in 94-98% of OPV2-naïve infants (aged 18-22 weeks) 2 weeks after mOPV2 challenge (23) and 82% of infants 2 weeks after one dose of nOPV2 (8). Further research is necessary to understand temporal differences in the induction of mucosal immunity to nOPV2 received at birth versus in later infancy.

The absence of positive PV2-specific neutralizing activity in stool in half the nOPV2 recipients after the birth dose warrants consideration. The lack of birth dose neutralization responses and the delayed peak in antibodies compared to older infants may reflect lower replication of PV in the neonatal gut or interference of maternal antibodies (serum IgG passively transferred through the placenta and secretory IgA in the breastmilk). While critical to protecting newborns, maternal antibodies are known to interfere with early vaccine responses (18,24), including those to poliovirus vaccines (25–27). High maternal antibodies measured in cord blood, in serum, or in colostrum have been associated with lower responses to a birth dose of oral poliovirus vaccine as measured in virus excretion in stool or serum antibody response (17,26–28). In our study, high serum neutralization at baseline was associated with lower odds of detectable neutralizing activity in stool at 2 weeks, but not at 4 weeks. Furthermore, we hypothesize that the low levels of type 2-specific IgA observed in the vaccine and placebo arms at baseline may be breastmilk-derived. However, clear differences in the type 2-specific IgA levels emerged from two weeks onwards after the first dose of nOPV2.

Alternatively, immunologic priming by nOPV2 birth dose might explain why we observed similar functional activity and binding antibody levels between birth dose responders and non-responders at 8 weeks. It has been suggested that priming may occur (via B cell memory) based on observations of strong and rapid antibody responses after a second vaccine dose in early life (18,29). However, our findings are not consistent with an amnestic response, as we observed similar magnitudes of neutralizing activity 2 weeks after both the first dose in responders and the second dose in non-responders. Additionally, only 3.9% of birth dose non-responders had detectable virus shedding after the first dose, while 92.2% shed after the second dose, suggesting that the vaccine might have initially failed to “take” due to inefficient replication of nOPV2. Notably, the vaccine virus shedding titers in stool (measurable by cell culture) at day 14 were very low in the newborn, as reported by the parent study (9), although previous study in older infants suggest that shedding peaks 7 days after vaccination (7). While mucosal immunity in the neonatal intestine typically develops rapidly after birth, concurrent with the gut microbiota (30), failure to induce detectable enteric IgA after one dose of nOPV2 in some newborns may also be explained by an age-dependent lack of endogenous IgA production in the intestine at birth (31).

We believe nOPV2 administration during the neonatal period may be a strategy to enhance population-level intestinal mucosal immunity in cVDPV2 outbreak settings. Neonatal vaccination offers several potential advantages: (i) logistically, infants have increased contact with the healthcare system near the time of birth, (ii) early vaccination could mitigate risks of a susceptibility window as maternal antibodies wane and before routine immunization, (iii) potentially, lower shedding of vaccine virus decreases risks of circulating virus, (iv) newborns are less likely to be exposed to other enteric infections known to diminish the effectiveness of OPV in older infants (24,32), and (v) newborns are less likely to develop vaccine-associated paralytic poliomyelitis (VAPP) because of maternally protective serum antibody (21). This trial provides preliminary evidence that the magnitude of intestinal mucosal neutralizing activity induced by neonatal nOPV2 administration is similar or higher to those observed with mOPV2 or nOPV2 challenges in later life. Further research on neonatal poliovirus vaccination is warranted to understand the optimal number and timing of doses, the duration of induced intestinal immunity, and to identify any contraindications or effect modification by factors such as prematurity.

## Data Availability

All data produced in the present study are available upon reasonable request to the authors

## Acknowledgments

Published as part of the ongoing efforts of the Dartmouth International Vaccine Initiative. The monovalent inactivated poliovirus vaccines used in the Luminex assays were provided by Bilthoven Biologicals (The Netherlands).

The International Centre for Diarrhoeal Disease Research, Bangladesh, acknowledges with gratitude the Bill and Melinda Gates Foundation (BMGF) for supporting this research and the Governments of Bangladesh and Canada for providing core and unrestricted support.

This activity was reviewed by the CDC and was conducted consistent with applicable federal law and CDC policy (e.g., 45 C.F.R. part 46, 21 C.F.R. part 56; 42 U.S.C. §241(d); 5 U.S.C. §552a; 44 U.S.C. §3501 et seq.). The findings in this article are those of the authors and do not necessarily represent the official position of the US Centers for Disease Control and Prevention.

---

**Table S1.** Baseline characteristics of the study participants. Abbreviations: nOPV2, novel oral polio vaccine type 2; Sd, standard deviation; BCG, Bacillus Calmette-Guérin. * All breastfed newborns were reported to be exclusively breastfed at birth, and partially breastfed at week 4

|  | **nOPV2 (N=110)** | **Placebo (N=105)** |
| --- | --- | --- |
|  | n/N (%) \| mean (sd) | n/N (%) \| mean (sd) |
| **Sex** |  |  |
| Male | 53/110 (48.2%) | 54/105 (51.4%) |
| Female | 57/110 (51.8%) | 51/105 (48.6%) |
| **Age (days)** | 1.1 (0.7) | 1.1 (0.6) |
| **Weight (kg)** | 2.9 (0.3) | 2.8 (0.3) |
| **Breastfeeding\*** |  |  |
| At baseline | 109/110 (99.1%) | 105/105 (100%) |
| At week 4 visit | 109/110 (99.1%) | 104/105 (99.1%) |
| **Received BCG vaccine** |  |  |
| At birth | 44/110 (40.0%) | 49/105 (46.7%) |
| Between birth and week 4 visit | 26/110 (23.6%) | 21/105 (20.0%) |

**Table S2.** Baseline characteristics of the responders and non-responders to the first dose of nOPV2 vaccine. *P* values are from *t*-tests and Pearson’s chi-squared tests. Responders are participants with stool neutralization titers ≥16 at week 2 and/or week 4. Abbreviations: nOPV2, novel oral polio vaccine type 2; Sd, standard deviation; BCG, Bacillus Calmette-Guérin.

|  | **Birth dose responder**<br>**(N=59)** | **Birth dose non-**<br>**responder (N=51)** |  |
| --- | --- | --- | --- |
|  | n/N(%) \| mean (sd) | n/N(%) \| mean (sd) | ***P* value** |
| **Sex male** | 26/59 (44.1%) | 27/51 (53.0%) | 0.35 |
| **Age (days)** | 1.2 (0.7) | 0.9 (0.7) | **0.04** |
| **Weight (kg)** | 2.9 (0.3) | 3.0 (0.4) | 0.20 |
| **Breastfeeding** | 59/59 (100%) | 50/51 (98.0%) | 0.28 |
| **Received BCG vaccine** | 24/59 (40.7%) | 20/51 (39.2%) | 0.88 |

**Figure S1.**
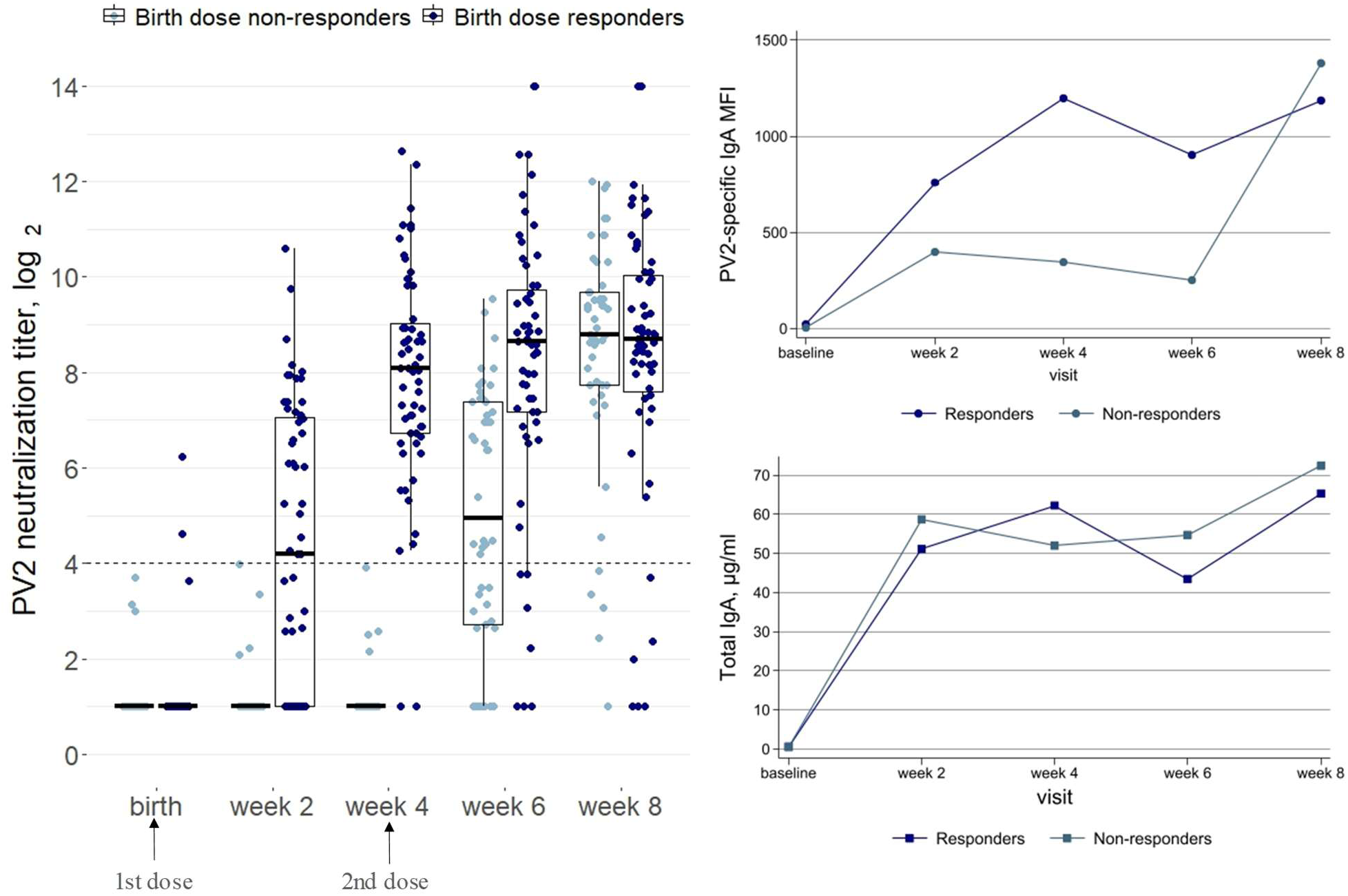
Distribution of poliovirus type 2 (PV2)-specific log_2_ neutralization titers, median total immunoglobulin A (IgA, µg/ml), and PV2-specific median fluorescence intensities (MFIs) IgA in stool samples after vaccination with the novel oral polio vaccine type 2 (nOPV2) in birth dose responders and non-responders. Responders are participants with stool neutralization titers ≥16 at week 2 and/or week 4. On the left plot: the dashed lines indicate the limit of positivity for the neutralization (i.e., titers ≥16); horizontal lines indicate the median; box and whisker indicate the interquartile range (IQR) and 1.5*IQR.

**Table S3.** Association of poliovirus type 2 (PV2)-specific detectable neutralization titers in stool two and four weeks after receiving the first dose of novel type 2 oral poliovirus vaccine (nOPV2) with the magnitude of PV2-specific neutralization in serum at baseline. *P* values are from Pearson’s chi-squared tests. Neutralization titers >4 were considered detectable. Abbreviations: OR, odds ratio; 95% CI, 95% confidence interval.

|  | Detectable neutralization titers in stool<br>2 weeks after 1<sup>st</sup> dose of nOPV2<br>(N=42/110) |  |  |  | Detectable neutralization titers in stool<br>4 weeks after 1<sup>st</sup> dose of nOPV2<br>(N=61/110) |  |  |  |
| --- | --- | --- | --- | --- | --- | --- | --- | --- |
|  | n/N (%) | OR | 95% CI | *P* | n/N (%) | OR | 95% CI | *P* |
| Baseline log<sub>2</sub><br>neutralization titers<br>in serum |  |  |  |  |  |  |  |  |
| High (6.8-10.5) | 8/35 (22.9%) | Ref | Ref | -- | 19/35 (54.3%) | Ref | Ref | -- |
| Medium (5.1-6.5) | 12/34 (35.3%) | 1.8 | 0.6 - 5.3 | 0.26 | 16/34 (47.1%) | 0.7 | 0.3 - 1.9 | 0.55 |
| Low (2.5-4.8) | 22/41 (53.7%) | 3.9 | 1.4 - 10.6 | **0.008** | 26/41 (63.4%) | 1.5 | 0.6 - 3.7 | 0.42 |

**Table S4.**
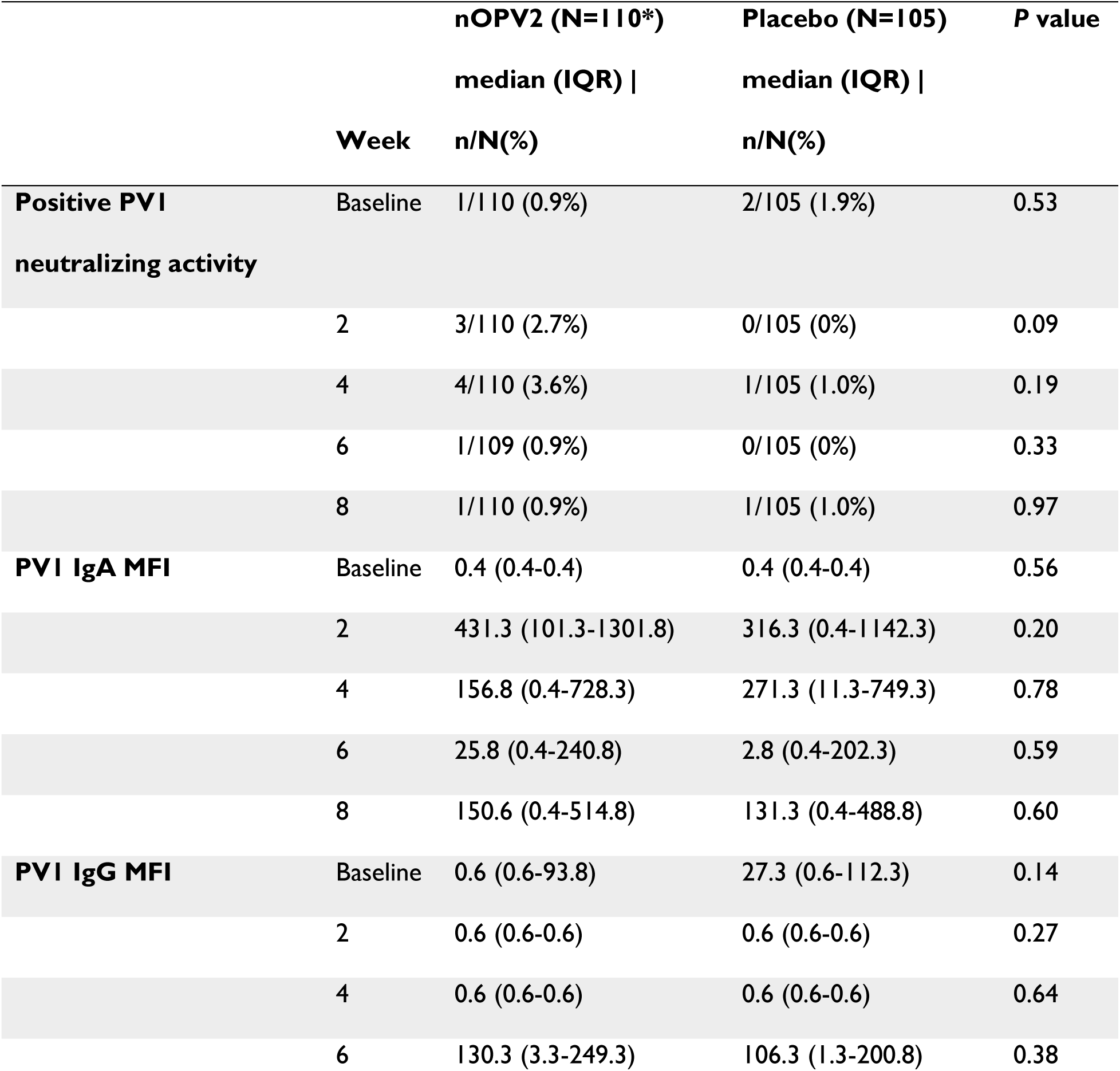
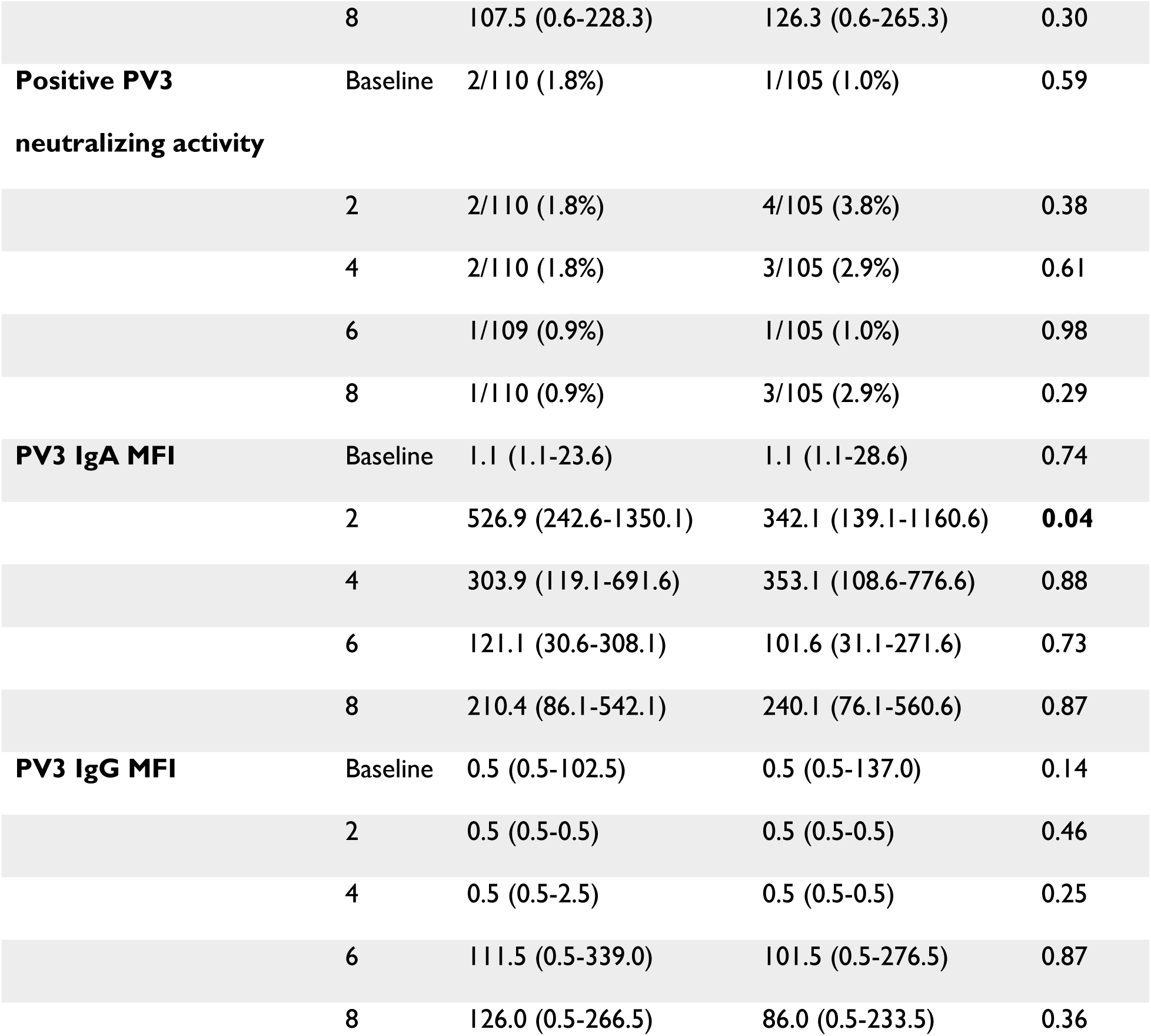
Poliovirus type 1 (PV1)- and type 3 (PV3)–specific intestinal mucosal responses following administration of novel type 2 oral poliovirus vaccine (nOPV2) or a placebo. *P* values are from Pearson’s chi-squared or Mann-Whitney U tests. Neutralization titers ≥16 were considered a positive neutralizing activity. Abbreviations: IgA, Immunoglobulin A; MFI, median fluorescence intensity; IQR, interquartile range. *n=109 in week 6 in the nOPV2 group.

**Table S5.** Poliovirus type 2 (PV2)–specific intestinal detectable neutralization titers following administration of novel type 2 oral poliovirus vaccine (nOPV2) or placebo. *P* values are from Pearson’s chi-squared tests. Neutralization titers >4 were considered detectable.

|  | **Week** | **nOPV2 (N=110)** | **Placebo (N=105)** | ***P* value** |
| --- | --- | --- | --- | --- |
|  |  | **n/N (%)** | **n/N (%)** |  |
| **Detectable PV2 neutralization titers** | Baseline | 6/110 (5.5%) | 7/105 (6.7%) | 0.71 |
|  | 2 | 42/110 (38.2%) | 17/105 (16.2%) | **<0.0001** |
|  | 4 | 61/110 (55.5%) | 13/105 (12.4%) | **<0.0001** |
|  | 6 | 96/109 (88.1%) | 20/105 (19.1%) | **<0.0001** |
|  | 8 | 106/110 (96.4%) | 17/105 (16.2%) | **<0.0001** |

**Table S6.** Poliovirus type2 (PV2)–specific intestinal mucosal responses following administration of novel type 2 oral poliovirus vaccine (nOPV2) stratified by i/ sex, ii/ BCG received at birth. *P* values are from Pearson’s chi-squared or Mann-Whitney U tests. Neutralization titers ≥16 were considered a positive neutralizing activity. Abbreviations: BCG, Bacillus Calmette-Guérin; IgA, Immunoglobulin A; MFI, median fluorescence intensity; IQR, interquartile range. *n=53 week 6 ** n=65 week 6

|  | Week | Male<br>(N=53*)<br>median (IQR)<br>\| n/N (%) | Female<br>(N=57)<br>median (IQR) \|<br>n/N (%) | *P* value | BCG at birth<br>(N=44)<br>median (IQR)<br>\| n/N (%) | No BCG at<br>birth (N=66**)<br>median (IQR) \|<br>n/N (%) | *P* value |
| --- | --- | --- | --- | --- | --- | --- | --- |
| **Positive PV2**<br>**neutralizing activity** | Baseline | 1/53 (1.9%) | 1/57 (1.8%) | 0.96 | 0/44 (0%) | 2/66 (3.0%) | 0.24 |
|  | 2 | 14/53 (26.4%) | 17/57 (29.8%) | 0.70 | 14/44 (31.8%) | 17/66 (25.8%) | 0.49 |
|  | 4 | 25/53 (47.2%) | 32/57 (56.1%) | 0.35 | 24/44 (54.6%) | 33/66 (50.0%) | 0.64 |
|  | 6 | 39/52 (75.0%) | 44/57 (77.2%) | 0.78 | 33/44 (75.0%) | 50/65 (76.9%) | 0.82 |
|  | 8 | 48/53 (90.6%) | 51/57 (89.5%) | 0.85 | 40/44 (90.9%) | 59/66 (89.4%) | 0.80 |
| **PV2 neutralization titer** | Baseline | 2 (2-2) | 2 (2-2) | 0.95 | 2 (2-2) | 2 (2-2) | 0.66 |
|  | 2 | 2 (2-19.2) | 2 (2-38.1) | 0.83 | 2 (2-38.1) | 2 (2-18.3) | 0.43 |
|  | 4 | 5.9 (2-400.7) | 40.0 (2-261.4) | 0.83 | 22.8 (2-406.4) | 26.1 (2-261.4) | 0.76 |
|  | 6 | 158.1 (16.9-459.4) | 174.2 (21.2-420.9) | 0.99 | 170.0 (16.9-482.8) | 165.9 (21.2-400.7) | 0.87 |
|  | 8 | 442.0 (248.9-1336.1) | 406.4 (174.2-731.7) | 0.11 | 412.6 (170.4-949.2) | 442.0 (211.9-995.7) | 0.53 |
| **PV2 IgA MFI** | Baseline | 9.6 (0.1-48.1) | 8.1 (0.1-52.1) | 0.94 | 14.6 (0.1-60.1) | 7.9 (0.1-48.1) | 0.33 |
|  | 2 | 647.1 (280.6-1753.1) | 655.1 (300.6-1379.1) | 0.74 | 673.6 (314.6-1565.4) | 651.1 (280.6-1409.1) | 0.71 |
|  | 4 | 737.1 (228.6-1417.6) | 682.1 (234.4-1608.4) | 0.82 | 786.6 (207.1-1417.6) | 691.4 (263.1-1979.6) | 0.53 |
|  | 6 | 520.1 (245.6-1309.6) | 573.1 (208.1-956.1) | 0.80 | 496.4 (173.9-1100.4) | 561.1 (255.6-1209.6) | 0.47 |
|  | 8 | 1187.1 (842.1-2421.6) | 1441.6 (625.6-2021.1) | 0.67 | 1179.1 (597.4-2293.4) | 1290.6 (842.1-1957.1) | 0.94 |

**Figure S2.**
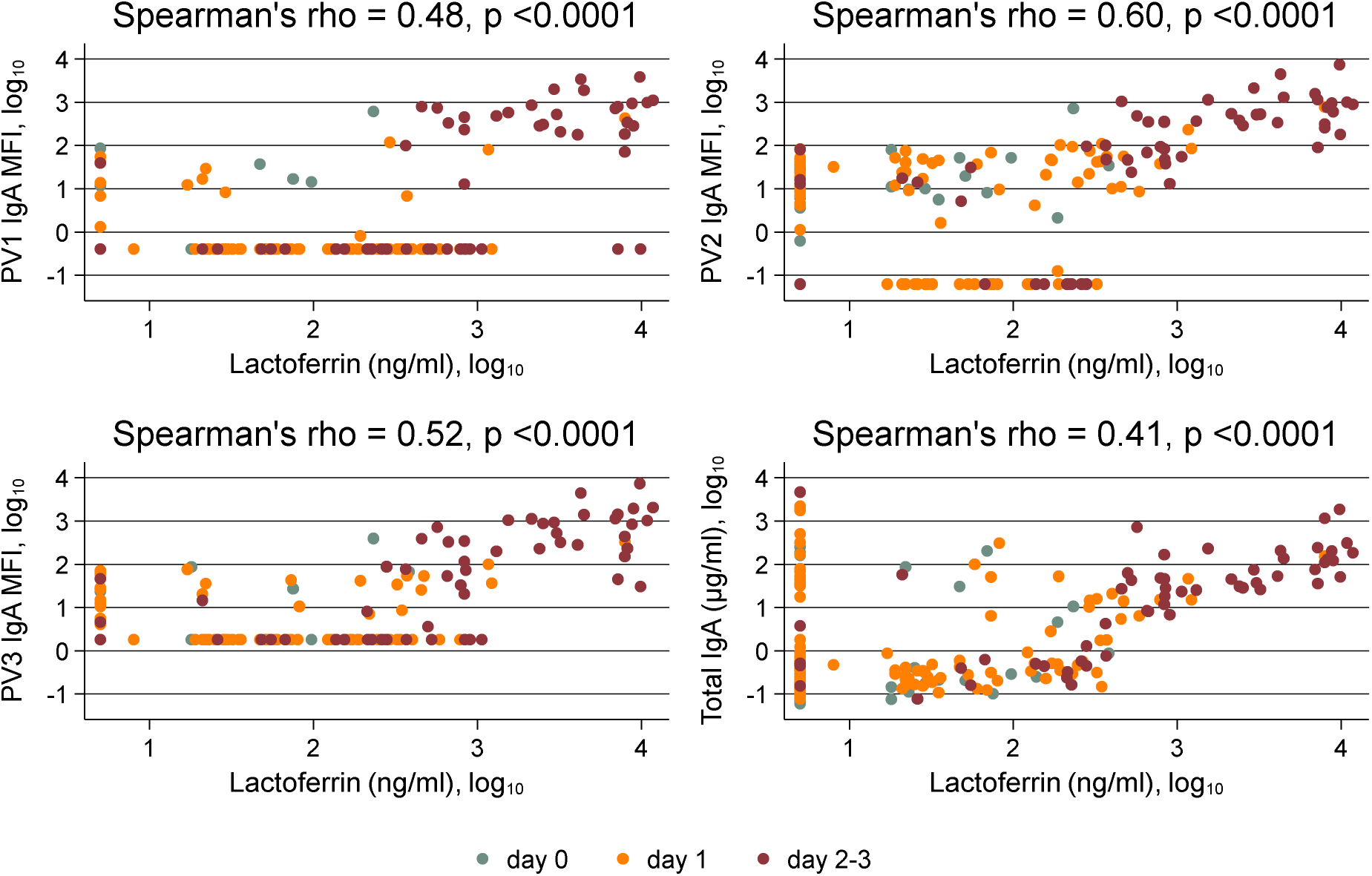
Correlation of total immunoglobulin A (IgA, µg/ml), poliovirus serotype-specific (PV1, PV2, PV3) median fluorescence intensities (MFIs) IgA, and lactoferrin levels (ng/ml) in stool at baseline (i.e., days 0-3; before vaccination) stratified by age at study enrollment (in days). Colors indicate the age at enrollment. Both randomization groups were included.

